# LuftiBus in the school (LUIS): a population-based study on respiratory health in schoolchildren

**DOI:** 10.1101/2020.10.14.20212076

**Authors:** Rebeca Mozun, Claudia E. Kuehni, Eva S. L. Pedersen, Myrofora Goutaki, Johanna M. Kurz, Kees de Hoogh, Jakob Usemann, Florian Singer, Philipp Latzin, Alexander Moeller

## Abstract

Respiratory disease is common in children and strongly associated with lifestyle and environmental exposures. Thus, it is important to study the epidemiology locally. *LuftiBus in the school* (LUIS) was set up to assess the respiratory health of schoolchildren in the canton of Zurich, Switzerland.

LUIS is a cross-sectional population-based study that was carried out 2013 to 2016. Children aged 6-17 years living in the canton of Zurich were eligible to participate. All schools in the canton were approached and the school head decided whether the school would participate and with which classes. Consenting parents answered a standardized questionnaire at home and assenting children completed a shorter questionnaire by interview at school. Trained technicians measured children’s lung function including spirometry, double tracer gas single-breath washout (DTG-SBW) and fractional exhaled nitric oxide (FeNO). Address histories of participants were geocoded to be linked with area-based socioeconomic measures and environmental exposures like spatiotemporal air pollution estimates for specific time periods and locations. A subgroup was seen again 12 months later using the same procedures to collect longitudinal data.

The study included 3870 children at baseline and 655 at the one-year follow-up. Median age was 12.7 years; 281 (8%) had wheezed in the past year. At baseline we collected 3457 (89%) parental and 3546 (92%) children’s questionnaires, and 3393 (88%) FeNO, 3446 (89%) spirometry, and 1795 (46%) DTG-SBW measurements.

LUIS is a rich resource of health-related data, with information on lung function, environmental exposures and respiratory health on Swiss schoolchildren.

**Take home message:** *LuftiBus in the school* (LUIS) is a population-based study with detailed lung function data and rich information on respiratory health in Swiss schoolchildren.

## Introduction

Respiratory disease is common and can affect quality of life and school performance in children [1]. Symptoms and diseases result from complex interactions between genetic factors and modifiable behavioural and environmental influences such as physical activity, passive and active smoking and air pollution [2]. The prevalence of respiratory symptoms such as wheeze and cough varies widely between countries, so local studies are important [3, 4]. Geographic differences in prevalence of respiratory diseases may result from differences in lifestyle, health care, socioeconomic factors, and environmental exposures, which has been the focus of many studies [2, 3, 5-7].

Previous research on the respiratory health of Swiss children focused on self-reported lower respiratory symptoms and farming environment [8-14]. None of the recent population-based studies included a detailed assessment of lung function and airway inflammation. Different examinations measure diverse aspects of lung physiology, such as airway inflammation, lung volumes, flows representing function of larger airways, and ventilation inhomogeneity representing the ventilation in peripheral airways. The same symptom might be caused by different underlying mechanisms, and similar underlying mechanisms may cause diverse symptoms in children. Thus, joint evaluation of results from different examinations and of reported symptoms would improve understanding of this complexity. The objectives of LUIS were to assess frequency and risk factors of upper and lower respiratory symptoms and to study associations between symptoms, different lung function assessments and environmental exposures. This article explains the methodology of LUIS and presents first results.

## Methods

### Study design and setting

Throughout the manuscript we use past tense for procedures or analyses that were done in the past and use present or future tense for those that are underway or planned. LUIS is a cross-sectional study that was conducted from November 2013 to December 2016 among schoolchildren aged 6 to 17 years living in the canton of Zurich (ClinicalTrials.gov: NCT03659838). A subsample of children was seen again one year later for collection of longitudinal data. The canton of Zurich is located in the north-eastern part of Switzerland and is the most populated canton in the country with over 1.5 million inhabitants, an 18% of the Swiss population [15].

### Study procedures

LUIS was embedded in a respiratory health promotion activity offered by *Lunge Zürich* [16-18]. *Lunge Zürich* is a non-profit organization that promotes respiratory health prevention and research [16]. LUIS used a special bus from *Lunge Zürich*, the *LuftiBus*, to visit schools within the canton of Zurich, further details on the *LuftiBus* are described in the online material [16]. All school heads in the canton of Zurich were invited to participate (Figure 1). They decided whether to take part in the project and with which classes. The study coordinator provided the schools with closed envelopes containing study information for parents and for children, an informed consent form and a questionnaire for parents. Teachers gave these envelopes to the students two weeks before the visit. Parents were asked to complete the questionnaire. Two study field workers, who were trained lung function technicians, visited each school with the study bus, the *LuftiBus* [16]. They collected the parental questionnaires and consent forms from the teachers and asked all students for oral and where appropriate written consent. The study bus contained computers and equipment for lung function testing. One field worker measured the children’s height and weight and performed the lung function tests, while the other interviewed the children using a short electronic questionnaire.

**Figure 1:**
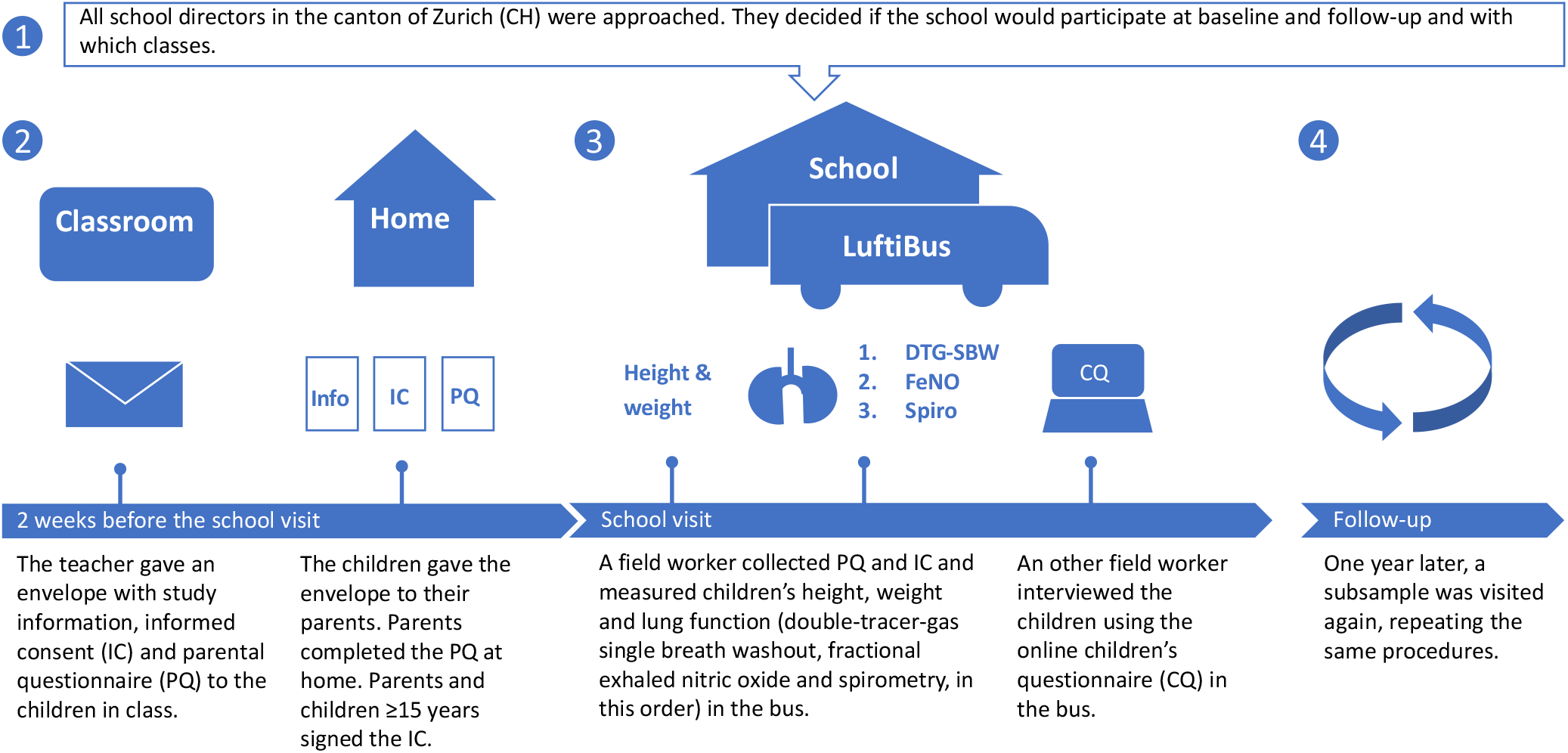
Design of the *LuftiBus in the school* (LUIS) study

Where the heads of the participating schools agreed, the school was visited again one year later (2015-2016) following the same procedures and performing the same tests as in the baseline visit.

### Data collection

#### The questionnaires

The study used two questionnaires: one for parents and one for children. Both included questions on respiratory symptoms and diagnoses including the key questions from the International Study on Asthma and Allergies in Childhood study and additional questions from the Leicester Respiratory Cohort questionnaires [3, 7]. The questionnaire to parents included questions about frequency, duration, severity and triggers of upper and lower respiratory symptoms, doctors’ diagnosis of asthma, medication, environmental factors, household characteristics, family history of asthma, and current and past addresses (Table 1). We geocoded participant’s addresses using a reference file from the Swiss Federal Statistical Office (Neuchâtel) [15]. The data from the parental questionnaire was entered into an Epidata database (version 4.2.0.0, EpiData Association, Denmark) at the Institute for Social and Preventive Medicine (ISPM) in Bern.

**Table 1:** Details of data collected in the *LuftiBus in the school study* (LUIS).

| Data sources | Collected data |
| --- | --- |
| **1. Questionnaires** |  |
| *For parents* | Upper and lower respiratory symptoms in the past 12 months: colds, rhinitis, snoring, cough, wheeze, frequency of wheeze/asthma attacks, triggers of cough and wheeze [3, 7] |
|  | Asthma diagnosis and hay fever [3, 7] |
|  | Management of wheeze / asthma in the past 12 months: number of doctor visits; medication use including short and long acting beta 2 agonists, inhaled corticosteroids, leukotriene receptor antagonists, and antihistaminics |
|  | Household: Current and past address, living on which floor, type of cooking stove, presence of oven or chimney, living on a farm |
|  | Environment and lifestyle: paternal and maternal smoking indoors or outdoors and number of cigarettes smoked, physical activity of the child, type of commuting to school, sports outside school, pets at home |
|  | Family: country of birth of the child, country of origin of the parents, siblings, parental education, parental history of asthma and chronic cough |
| *For children* | Currently having a cold; currently coughing |
|  | Upper and lower respiratory symptoms in the past 12 months: colds, rhinitis, waking up with a dry mouth as a proxy for mouth breathing, cough, wheeze, triggers of cough and wheeze [3, 7] |
|  | Asthma diagnosis and hay fever [3, 7] |
|  | Medication during the day of the visit: inhaled short and long acting beta two agonists, inhaled corticosteroids, leukotriene receptor antagonists |
|  | Environment and lifestyle: frequency of active smoking of cigarettes, shishas and e-cigarettes; paternal and maternal smoking |
| **2. Anthropometrics** | Height and weight measured in the bus without wearing shoes<br>Body mass index z-scores calculated using World Health Organisation reference tables [32] |
| **3. Lung function measurements** |  |
| *Spirometry* | Masterlab, Jaeger, Würzburg, Germany. Mouthpiece, filter, nasal clamp<br>Main outcomes: forced expiratory flows and volumes [28] |
| *FeNO* | Fast response chemiluminescence analyzer CLD 88, Eco Medics AG, Duernten, Switzerland. Mouthpiece, filter, no nasal clamp<br>Main outcome: mean FeNO (ppb) [27, 33] |
| *DTG-SBW (He, SF6)* | Exhalyzer D, Eco Medics AG, Duernten, Switzerland. Mouthpiece, filter, nasal clamp [23]<br>Main outcome: phase III slope |
| **4. Air pollution** | We will use data from air pollution models developed by de Hoogh et al. [30, 31] to estimate participants' average exposure to NO<sub>2</sub> and PM<sub>2.5</sub> for specific time windows and locations, based on participants' exact address history. |
| **5. Swiss-SEP** | An area-based measure of socioeconomic status based on data about rent per square meter, education and occupation of households' heads, and household crowding, which ranges from 0 (lowest) to 100 (highest), developed as part of the Swiss National Cohort study [20, 21] |

FeNO: Fractional exhaled nitric oxide. ppb: parts per billion. DTG-SBW: double tracer gas single breath washout. He: helium. SF6: sulphur hexafluoride. NO<sub>2</sub>: nitrogen dioxide. PM<sub>2.5</sub>: particulate matter of aerodynamic diameter of 2.5µm or less.

Children replied to the questionnaire via interview by the field workers in the bus. It included questions about current respiratory health such as rhinitis, cough and wheeze, the presence of cough or a cold on the day of the school visit, inhaled or oral asthma medication on the day of the visit, active smoking, and parental smoking. To help the children correctly report asthma medications, the field workers used a poster with pictures and commercial names of all commonly used inhaled and oral medications in Switzerland (online Figure S1). Children’s answers were directly entered by field workers into the web-based Research Electronic Data Capture (REDCap) database [19].

#### Area-based index of socioeconomic position in Switzerland (Swiss-SEP)

The Swiss-SEP was developed by the Swiss National Cohort (SNC) as an area-based measure of socioeconomic status that incorporates information on rent per square meter, education and occupation of households’ heads, and number of persons per household room [20, 21]. The Swiss-SEP ranges from 0 (lowest) to 100 (highest). We estimated the Swiss-SEP for our participants by matching the geocodes of our participants addresses to the nearest geocode in the Swiss-SEP dataset. If the address of a participant was missing, we assigned the median Swiss-SEP among participants of the same school. In this manuscript we used a dataset from the SNC to assess the distribution of Swiss-SEP among households from the canton of Zurich with at least one child in school-age (6-17 years) and compared it to the distribution of Swiss-SEP of our study participants.

#### Lung function measurements

Children’s standing height and weight were measured without shoes and recorded in the spirometry database. Lung function tests were performed to children inside the bus in the following order: 1) double tracer gas single-breath washout (DTG-SBW), 2) fractional exhaled nitric oxide (FeNO), and 3) spirometry.

#### DTG-SBW

DTG-SBW was performed using a validated and commercially available setup (Exhalyzer D^®^, Eco Medics AG, Duernten, Switzerland) [22]. The patient interface consisted of a bacterial filter and a snorkel mouthpiece to prevent air leaks near the mouth. According to current paediatric equipment recommendations, a dead space reducer and a disposable hygienic insert provided by the manufacturer were used [23]. The DTG mixture contained 26.3% helium (He), 5% sulphur hexafluoride (SF6), 21% oxygen (O2), and balance dinitrogen (N2) from pressurized cylinders (Carbagas, Bern, Switzerland). Children were watching a video for distraction while breathing through an open bypass system. Directly before tracer gas wash-in, the apparatus dead space and the bias flow was flushed with double-tracer gas or 100% O2 during expiration to prevent re-inspiration of expired or ambient gas [24]. DTG-SBW was performed in accordance to the European Respiratory Society (ERS) consensus on inert gas washout testing [23]: after establishing natural tidal breathing for at least 5 breaths monitored by online flow-volume loops and tidal volumes varying less than 10%, the system was flushed with the DTG mixture. Within only one breath, the children inhaled the double-tracer gas and exhaled it back. SBW tests were done in triplicate. After each test, at least 10 breaths at room air followed until the online monitoring of the gas signals returned to baseline [23]. Primary outcome of the DTG-SBW was the mean slope from the tidal (alveolar) phase III from three technically acceptable DTG-SBW curves, normalized for tidal volume (SnIII) [25]. The magnitude of phase III slopes from He and SF6 relates to acinar branching asymmetry and small airways obstruction [26]. DTG-SBW is a fast and easy to perform test with low variability and high repeatability [22].

#### FeNO

FeNO was measured with a single breath on-line method according to recommendations [27]. We used a fast response chemiluminescence analyser CLD 88, Eco Medics AG, Duernten, Switzerland. The field workers calibrated the FeNO device every morning and the gas was calibrated once per month. Children breathed air free of nitric oxide via the Denox system (EcoMedics, Dürnten). This setup also included a bacterial filter and disposable hygienic mouthpiece. After ten breaths, children exhaled against an adjusted expiratory resistance with a constant target flow of 50ml/sec for at least 4 seconds for children younger than 12 years and 6 seconds for children aged 12 years or older. We aimed for at least two reproducible exhalations with the nitric oxide plateau values within 10% of each other. Results were stored digitally, including information on FeNO (parts per billion, ppb), expiratory time (seconds), duration of the plateau (seconds), and average flow in the plateau (ml/s). The primary outcome was FeNO.

#### Spirometry

Spirometry was performed using Masterlab, Jaeger, Würzburg, Germany according to American Thoracic Society (ATS) / ERS recommendations [28]. The children performed 3 to 5 forced expiratory manoeuvres. Spirometry parameters were recorded and stored digitally using the Sentry Suite software, Carefusion, Hoechberg, Germany. Spirometry parameters included forced vital capacity (FVC), forced expiratory volume in the first second (FEV1), peak expiratory flow (PEF), FEV1/FVC ratio, maximum expiratory flow at 75%, 50% and at 25% FVC (FEF75, FEF50, FEF25) and between the 25% and 75% of FVC (FEF25–75). The spirometry flow-volume curve of the best effort was printed on paper. Primary outcomes were forced expiratory flows and volumes. Absolute values of spirometry parameters were expressed as z-scores according to Global Lung Initiative reference values [29].

#### Air pollution assessment

Spatiotemporal data on nitrogen dioxide (NO2) and particulate matter with an aerodynamic diameter of 2.5µm or less (PM2.5) will be obtained from models developed by de Hoogh et al. [30, 31]. Daily average NO2 concentrations were modelled across Switzerland using a multistage framework with mixed-effect and random forest models at a fine spatial resolution of 100×100m [30] combining the Ozone Monitoring Instrument (OMI) NO2 product with Copernicus Atmosphere Monitoring Service (CAMS), land use and meteorological variables. Daily PM2.5 concentrations across Switzerland were estimated first by a combination of a mixed effect model and a generalised additive mixed model at a 1×1km resolution using Multiangle Implementation of Atmospheric Correction (MAIAC), spectral aerosol optical depth (AOD) data, and second by support vector machine algorithms used to predict precise exposure estimates at a 100×100m resolution with spatiotemporal predictor data [31]. These air pollution models allow to estimate participants’ average exposure to NO2 and PM2.5 for specific time windows and locations. We will link the geocoded residential and school addresses of study participants to NO2 and PM2.5 data by GIS (geographic information system) overlay.

### Ethics and data safety

The ethics committee of the canton of Zurich approved the study (KEK-ZH-Nr: 2014-0491). Parents signed the informed consent and completed a detailed questionnaire. Children assented orally and those aged 15 years or older did also sign. Participant identifiable information was stored in a separate database and only pseudoanonymized datasets are used for analysis. Paper-based data were securely locked at ISPM and only accessible to the study team. Raw data of DTG-SBW, FeNO and spirometry measurements were stored in a safe study server during recruitment and were thereafter encrypted and securely stored in the ISPM protected server. Access to the server and the study databases is restricted to members of the study team. Study datasets are securely stored and routinely backed-up in protected servers of ISPM Bern. Study collaborators and other researchers can obtain datasets for analysis if a detailed concept sheet is presented for the planned analyses and approved by the principal investigators (AM, PL and CK).

### Data preparation and analysis

We calculated body mass index (BMI) z-scores according to World Health Organisation references and categorised BMI z-scores ≥2 as obesity, ≥1 to <2 as overweight and ≤-2 as thinness [32]. In this manuscript, we compared participants’ characteristics between age groups using P values for a trend. We used the software STATA (Version 16.1, StataCorp., College Station, TX) for statistical analysis. In our planned analyses, we will select confounders a priori based on Directed Acyclic Graphs (DAGs). Results from DTG-SBW, FeNO or spirometry tests which did not meet the quality standards described in the online material will not be included in analyses. Details on data quality assessments can be found in the online material. We will adhere to STROBE reporting guidelines in manuscripts using LUIS datasets [34].

## Results

### Participating schools

All 490 schools in the canton of Zurich were invited to participate (online Table S1). At baseline 37 schools took part and 3870 participants consented. One year later, 19 schools were visited a second time and 655 children were followed-up. The canton of Zurich (Figure 2) had mostly schools in small (47%) and large urban areas (41%) and fewer in rural areas (12%) (Figure 3, online Table S1) [35, 36]. At baseline, LUIS visited 18 schools in small (49%) and 15 in large (40%) urban areas and 4 in rural areas (11%). At follow up, 13 schools in small (68%) and 4 in large (21%) urban areas and 2 in rural areas (11%) were visited. Median Swiss-SEP was 66.2 (interquartile range, IQR: 58.2-73.0) for households with at least one school-aged child in the canton of Zurich and 69.2 (IQR: 61.2-76.3) for LUIS participants at baseline and 69.4 (IQR: 64.7-74.8) at follow-up (Figure 4, Figure S2).

**Figure 2:**
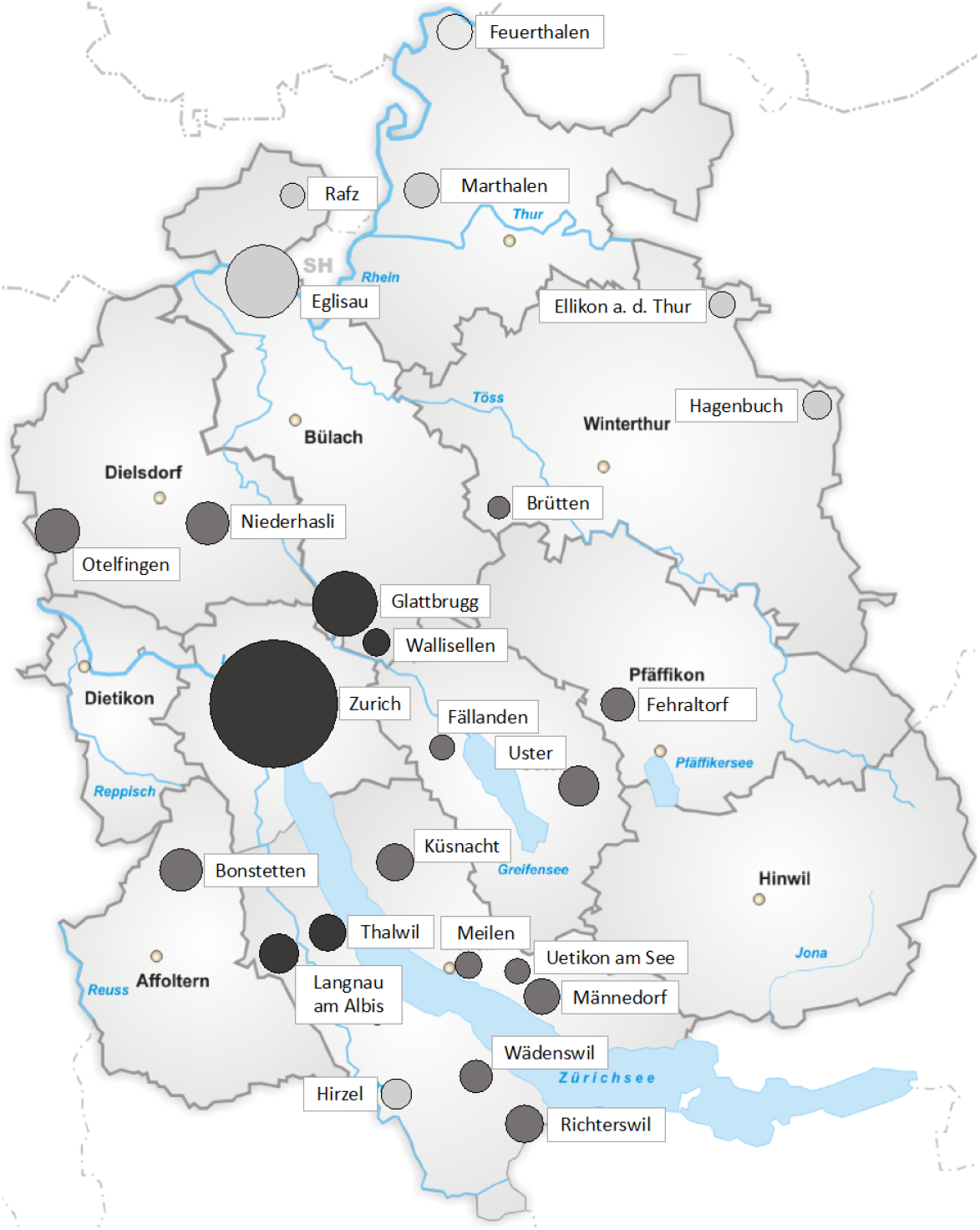
Areas from the canton of Zurich that were visited by the *LuftiBus in the school* (LUIS) study. The area of the circle is proportional to the number of participants per location. The colour of the circle indicates the degree of urbanization of the region (from darker to lighter grey: large urban, small urban, rural areas) according to the Swiss federal office of statistics classification. Definition of urbanization degree: Cities or large urban area: At least 50% lives in high-density clusters. Towns and suburbs or small urban area: Less than 50% of the population lives in rural grid cells and less than 50% lives in a high-density cluster. Rural area: More than 50% of the population lives in rural grid cells.

**Figure 3:**
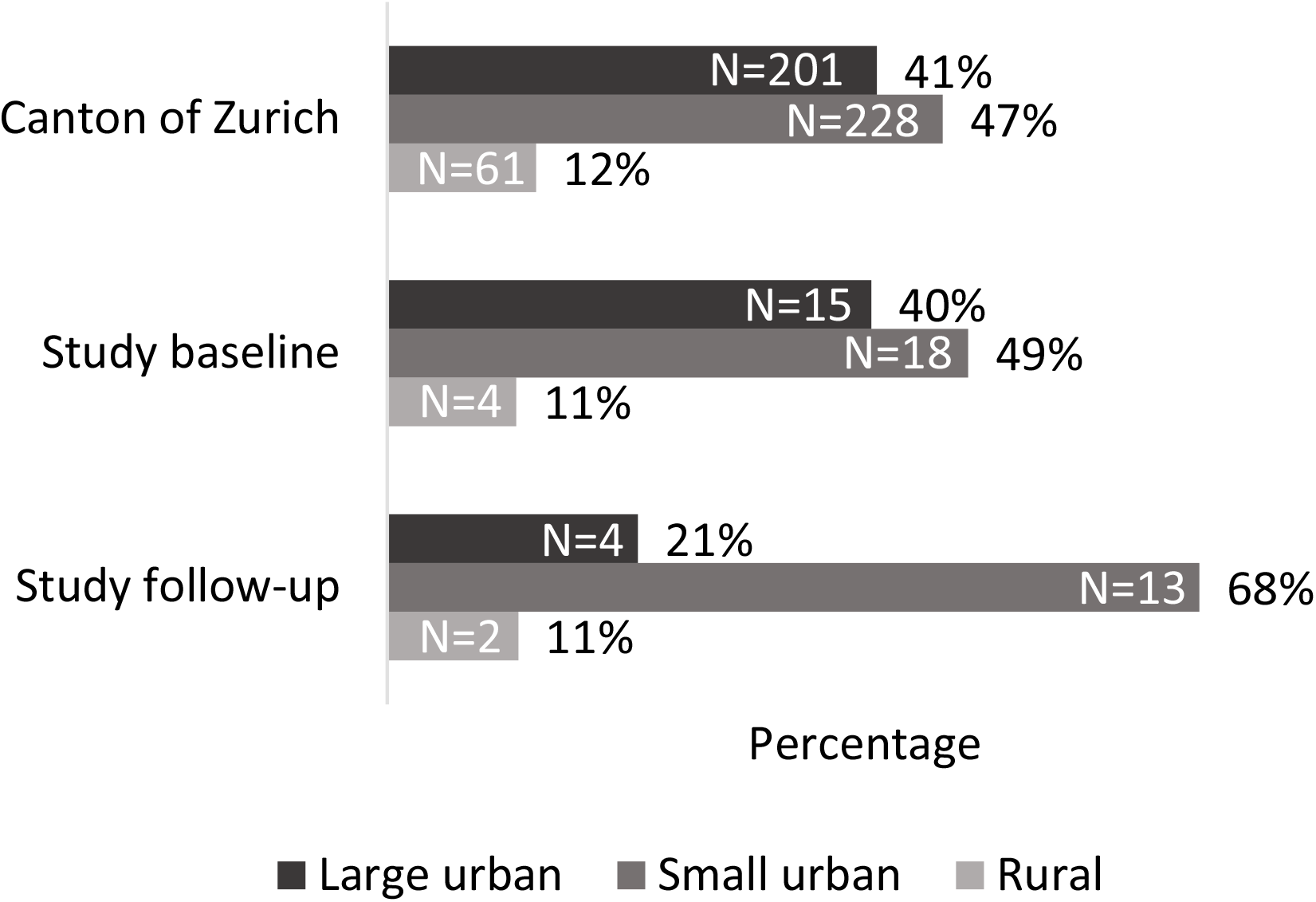
Proportion of schools by degree of urbanisation in the canton of Zurich and in the *LuftiBus in the school* (LUIS) study at baseline and follow-up. Definition of urbanization degree: Cities or large urban area: At least 50% lives in high-density clusters. Towns and suburbs or small urban area: Less than 50% of the population lives in rural grid cells and less than 50% lives in a high-density cluster. Rural area: More than 50% of the population lives in rural grid cells.

**Figure 4:**
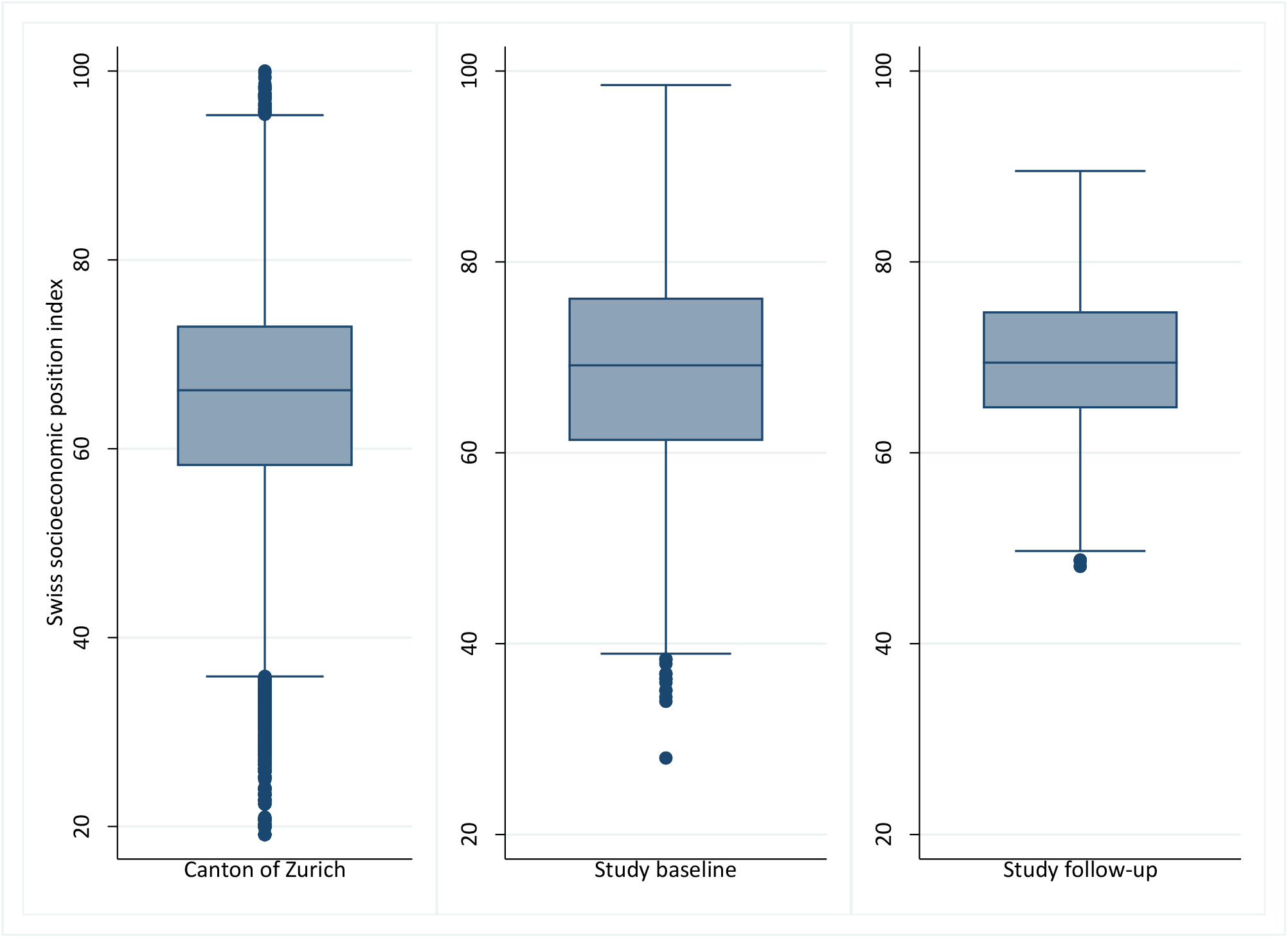
Box plots showing the distribution of Swiss socioeconomic position index (Swiss-SEP) for families with at least one child aged 6-17 years and living in the canton of Zurich and in the *LuftiBus in the school* (LUIS) study at baseline and follow-up. The median is represented by a horizontal line inside the box, 25 and 75 percentiles by the upper and lower box horizontal lines of the box, minimum and maximum values are shown as whiskers, and points outside the whiskers are outliers.

### Completeness of datasets

At baseline, 3457 participants (89%) provided parental questionnaires, 3546 (92%) answered the children’s questionnaire, 3446 (89%) did spirometry, 3393 (88%) FeNO and 1795 (46%) DTG-SBW tests (Table S2). At the one year follow-up, data from parental questionnaires was available for 629 children (96%), data from children’s questionnaires for 640 (98%), results from spirometry for 614 (94%), results from FeNO testing for 588 (90%) and results from DTG-SBW for 496 children (76%).

### Characteristics of the study population

LUIS included 987 children aged 6-9 years (2%), 1723 aged 10-13 years (45%) and 1160 aged 14-17 years (30%) at baseline (Table 2). Current wheeze was reported by parents of 281 (8%) children. Exercise induced wheeze was more common among older children, and night cough among younger children. Use of inhaled short acting beta two agonists (SABA) and inhaled corticosteroid (ICS) in the past year was reported by parents of 257 (8%) and 179 (5%) children, respectively. Parents of 1108 (32%) reported that their child had a high physical activity level. This was more common for younger children. Mothers of 574 (17%) and fathers of 758 (22%) children reported to smoke. 580 children (15%) were overweight and 194 (5%) obese, with obesity being more common in older children.

**Table 2:** *LuftiBus in the school study* (LUIS): Characteristics of the study participants.

|  | Total<br>N=3870 | Age groups (years) |  |  | P trend |
| --- | --- | --- | --- | --- | --- |
|  |  | 6 to 9<br>N=987 | 10 to 13<br>N=1723 | 14 to 17<br>N=1160 |  |
| Male sex | 1937 (50%) | 509 (52%) | 842 (49%) | 586 (51%) | - |
| Age in years, mean (SD) | 12.1 (2.7) | 8.2 (1.0) | 12.4 (1.2) | 15.0 (0.7) | - |
| Body mass index |  |  |  |  |  |
| Thinness (z-score ≤ -2) | 139 (4%) | 34 (4%) | 76 (4%) | 29 (3%) | 0.002 |
| Normal weight (z-score > -2, < 1) | 2850 (76%) | 752 (80%) | 1251 (74%) | 847 (75%) |  |
| Overweight (z-score ≥ 1, < 2) | 580 (15%) | 126 (13%) | 276 (16%) | 178 (16%) |  |
| Obesity (z-score ≥ 2) | 194 (5%) | 33 (3%) | 93 (5%) | 68 (6%) |  |
| Level of physical activity |  |  |  |  |  |
| Low | 332 (10%) | 26 (3%) | 140 (9%) | 166 (18%) | <0.001 |
| Moderate | 1976 (58%) | 484 (51%) | 928 (60%) | 564 (61%) |  |
| High | 1108 (32%) | 443 (46%) | 475 (31%) | 190 (21%) |  |
| Practice of sports outside school | 2612 (76%) | 767 (81%) | 1216 (79%) | 629 (68%) | <0.001 |
| Symptoms in the past 12 months |  |  |  |  |  |
| Wheeze | 281 (8%) | 75 (8%) | 137 (9%) | 69 (8%) | 0.864 |
| Exercise-induced wheeze | 274 (8%) | 43 (5%) | 134 (9%) | 97 (11%) | <0.001 |
| Night cough apart from colds | 394 (12%) | 142 (15%) | 154 (10%) | 98 (11%) | 0.003 |
| Rhinitis apart from colds | 1049 (31%) | 217 (23%) | 501 (33%) | 331 (36%) | <0.001 |
| Habitual snoring (almost every night) | 163 (5%) | 55 (6%) | 60 (4%) | 48 (5%) | 0.584 |
| Use of inhaled medication |  |  |  |  |  |
| Short acting beta two agonists | 257 (8%) | 63 (7%) | 123 (8%) | 71 (8%) | 0.344 |
| Inhaled corticosteroids | 179 (5%) | 36 (4%) | 90 (6%) | 53 (6%) | 0.050 |
| Maternal smoking | 574 (17%) | 121 (13%) | 266 (17%) | 187 (20%) | <0.001 |
| Paternal smoking | 758 (22%) | 171 (18%) | 335 (22%) | 252 (27%) | <0.001 |
| Pets in the household | 1469 (43%) | 313 (33%) | 720 (47%) | 436 (47%) | <0.001 |
| Living in a farm | 85 (2%) | 12 (1%) | 46 (3%) | 27 (3%) | 0.021 |

Information obtained from the parental questionnaires at baseline. P trend: p value for a trend across ordered categories of age groups.

## Discussion

LUIS is a large school-based study performed 2013-2016 in the canton of Zurich, Switzerland which comprised a thorough assessment of respiratory symptoms, lung function, lifestyle and environmental exposures.

### Strengths and weaknesses

The main strength of LUIS is the wealth of health data that were collected, which includes three different lung function tests: spirometry, FeNO measured using high resolution equipment and the novel DTG-SBW. This permits the investigation of different physiological aspects of lung health. A subsample of participants repeated lung function measurements a year later, which allows to assess lung growth. We collected detailed questionnaire data on respiratory symptoms, diagnoses and lifestyle both from parents and from children, which allows comparisons between their answers. Information was collected on lower respiratory symptoms like wheeze and chronic cough, and on upper respiratory symptoms such as rhinitis, hay fever and snoring. Respiratory symptoms are influenced by seasonality but the *LuftiBus* visited the schools throughout different seasons, which gives the possibility to take seasonality into account in our analyses. Participants’ address histories enable us to use state-of-the-art methods [30, 31] to estimate air pollution exposure during specific time windows.

The main weakness is that it was not a perfectly random sample of schoolchildren. Participation was decided by the heads of the schools, and classes were recruited as a whole. We do not have individual data on non-participating students, which limits the possibilities to assess representativeness of the sampled population. However, the distribution of participating schools in rural and urban areas at baseline was similar to the average distribution of schools in the canton of Zurich. At follow-up, schools from small urban areas were slightly overrepresented in our study. The area-based socioeconomic index of participating families was roughly comparable to that of all households with school-aged children from the canton of Zurich. Thus, we think LUIS gives a fair picture of the school-aged population from the canton of Zurich. Another limitation is the lack of objective measurements of atopy such as skin prick tests or serum IgE levels. This limits the interpretation of FeNO data. We had however proxy measures of atopy i.e. reported hay fever and atopic dermatitis.

### Outlook

LUIS will cover knowledge gaps on children’s respiratory health. This dataset allows to determine prevalence of upper and lower respiratory symptoms, to describe lifestyle characteristics like physical activity and smoking [37], and to study associations with lung function. We are able to use information from both parents and children and several lung function tests including markers of ventilatory function (spirometry), ventilation inhomogeneity (DTG-SBW) and Th2 airway inflammation (FeNO), to answer research questions in a comprehensive way [33, 38]. For example, DTG-SBW data allows to detect even subtle changes in peripheral airways, that may not be identified by spirometry alone [38]. The broad set of data collected allows us to describe lung function in healthy children, and to report normal values. The LUIS study makes it also possible to study effects of air pollution on respiratory symptoms and lung function.

In conclusion, *LuftiBus in the school* (LUIS) provides a unique opportunity to assess the respiratory health in Swiss schoolchildren, and its association with health behaviours and air pollution. Better understanding of factors that influence Swiss schoolchildren’s respiratory health may help to establish new recommendations and influence policy makers’ decisions.

## Data Availability

Study collaborators and other researchers can obtain datasets for analysis if a detailed concept sheet is presented for the planned analyses and approved by the principal investigators (AM, PL and CK).

## The LuftiBus in the school study group

The study PIs Prof. Alexander Moeller, Prof. Claudia Kuehni and Prof. Philipp Latzin conceptualised and designed the study. Alexander Moeller supervised data collection and was responsible for the overall conduct of the study. Philipp Latzin was responsible for the lung function measurements. Claudia Kuehni was responsible for the questionnaire design and data analysis. Rebeca Mozun was responsible for data management and coordination. Florian Singer and Johanna Kurz are responsible for management and analysis of DTG-SBW data. Myrofora Goutaki, Eva S.L. Pedersen and Cristina Ardura-Garcia (Institute of Social and Preventive Medicine, University of Bern, Switzerland) support statistical analysis. Jakob Usemann, Martin Röösli (Swiss Tropical and Public Health Institute, Basel, Switzerland) and Kees de Hoogh are responsible for the air pollution data.

## Acknowledgements

We thank the school teams and the families for participating in the study. We thank the field workers and study personnel for conducting the study. We thank Andras Soti, Marc-Alexander Oestreich, Corin Willers (Paediatric Respiratory Medicine, Children’s University Hospital of Bern, University of Bern, Switzerland), Léonie Hüsler, Eugénie Collaud and Carmen C. M. de Jong (Institute of Social and Preventive Medicine, University of Bern, Switzerland) for their help in the assessment of the quality of the spirometry flow-volume curves. We thank Claudia Berlin and the Swiss National Cohort (SNC) (www.swissnationalcohort.ch) for providing data on the Swiss-SEP for the canton of Zurich. We thank the Swiss federal statistical office for providing data on schools and urbanisation degree of municipalities in the canton of Zurich.

## Authors contributions

Alexander Moeller, Claudia E. Kuehni and Philipp Latzin conceptualised and designed the study. Rebeca Mozun analysed the data and drafted the manuscript. Eva S.L. Pedersen supported the statistical analysis. All authors gave input for interpretation of the data. All authors critically revised and approved the manuscript.

