## Supplement for "LuftiBus in the school (LUIS): a population-based study on respiratory health in schoolchildren"

### Supplementary online material

**Table of contents:**

Additional methods……………………………………………………………………………………………………….2
Table S1: Characteristics of schools in the canton of Zurich and visited by the LuftiBus in the school study (LUIS) at baseline and one-year follow-up by degree of urbanization……………………………………………………………………………………………………….....8
Table S2: Number of participants in the LuftiBus in the school (LUIS) study with available information on parental questionnaires (PQ), children's questionnaires (CQ), spirometry (Spiro), and fractional exhaled nitric oxide (FeNO) at baseline and one-year follow-up…………………………………………………………………………………………………………………9
Figure S1: Poster of asthma medications used in the LuftiBus in the school (LUIS) study to help children answer which medication they had taken the day of the visit (in German)………………………………………………………………………………………………………………10
Figure S2: Histograms showing the distribution of the Swiss socioeconomic position index for families from the canton of Zurich with at least one child aged 6-17 years living in the household and in the LuftiBus in the school (LUIS) study at baseline and one year follow-up……………………………………………………………………………………………...11
Figure S3: Fit lines of double tracer gas single breath washout (DTG-SBW), differences in molar mass (MM) plotted against expired volume……………………………………………..12

Rebeca Mozun*[1,2], Claudia E. Kuehni*[1,3], Eva S. L. Pedersen[1], Myrofora Goutaki[1,3], Johanna M. Kurz[3], Kees de Hoogh[4,5], Jakob Usemann[6,7], Florian Singer[3,8], Philipp Latzin[#3], Alexander Moeller[#6]
*These authors contributed equally to the work
#These authors contributed equally to the work

[1]Institute of Social and Preventive Medicine, University of Bern, Switzerland
[2]Graduate School for Cellular and Biomedical Sciences, University of Bern, Switzerland
[3]Paediatric Respiratory Medicine, Children's University Hospital of Bern, University of Bern, Switzerland
[4]Swiss Tropical and Public Health Institute, Basel, Switzerland
[5]University of Basel, Basel, Switzerland
[6]Division of Respiratory Medicine, University Children's Hospital Zurich and Children's Research Center, University of Zurich, Switzerland
[7]University Children's Hospital Basel (UKBB), Basel, Switzerland
[8]PEDNET, paediatric clinical trial unit, Children's University Hospital of Bern, University of Bern, Switzerland

### Additional methods

LuftiBus in the school (LUIS) is a study funded by *Lunge Zürich,* a non-profit organization that promotes respiratory health prevention and research. LUIS used a special bus ("LuftiBus") to visit schools within the canton of Zurich. Inclusion criteria were consent to participate and age 6 to 17 years. There were no predefined exclusion criteria.

#### LuftiBus

LuftiBus is a broader project of *Lunge Zürich* for the prevention and early detection of lung diseases and has been ongoing for 30 years [1, 2]. LuftiBus offers lung function tests to adults of the general public living in the metropolitan region of Zurich at a non-profit cost of 10 CHF [3]. Previous research has used data from the LuftiBus lung function campaign to calculate reference equations for lung function [3, 4]. LuftiBus data was linked with data form the Swiss National Cohort [5] to assess time trends in the prevalence of airway obstruction among adults from the general population [6], occupational risk factors for airway obstruction [7], and spatial risk pattern of respiratory morbidity [8].

#### Data collection: double tracer gas single breath washout (DTG-SBW)

The gas mixture had the same molar mass (MM, g.mol-1) as medical-grade air, such that any detectable changes compared with normally expired molar mass can be attributed to relative changes in helium (He) and sulphur hexafluoride (SF6) concentrations [9]. MM was measured by a side-stream ultrasonic flowmeter, tidal flows by a main-stream ultrasonic flowmeter. On each test day, signal calibration and verification were performed prior to testing. The main-stream ultrasonic flowmeter was calibrated using a 1 L precision syringe. The side-stream ultrasonic flowmeter and the oxygen (O2) and carbon dioxide (CO2) sensors were calibrated using medical-grade calibration gas and pure O2 (Carbagas, Bern, Switzerland).

### Air pollution assessments

Individual air pollution exposure will be estimated for nitrogen dioxide (NO2) and for particulate matter with an aerodynamic diameter of 2.5µm or less (PM2.5) using air pollution models developed by de Hoogh et. al [10, 11]. Daily average NO2 concentrations from 2005 to 2016 were modelled in a multistage framework with mixed-effect and random forest models at a fine spatial resolution (100x100 m) [10]. The model incorporates spatial and temporal predictors including satellite-derived data from the Ozone Monitoring Instrument and Copernicus Atmosphere Monitoring Service, and road, land use, topography and meteorological information. Daily NO2 monitoring data were obtained from the Immisionsdatabank Luft (IDB Luft, FOEN, Bern, Switzerland) measured at 67 sites in 2005 and increased to 110 sites in 2016. The model was able to explain approximately 73% of the overall spatiotemporal variation in NO2 measurements [10].

Spatiotemporal resolved models were developed to predict daily PM2.5 exposure [11]. First, daily PM2.5 concentrations were estimated by a combination of a mixed effect model and a generalized additive mixed model at a 1×1km resolution across Switzerland using Multiangle Implementation of Atmospheric Correction (MAIAC) spectral aerosol optical depth (AOD) data. Second, support vector machine algorithms were used to predict precise exposure estimates at a 100 x 100m resolution in using spatiotemporal predictor data. The global (1 km) and local (100 m) models explain on average 73% of the total, 71% of the spatial and 75% of the temporal variation globally, and on average 89% of the total, 95% of the spatial and 88% of the temporal variation locally in measured PM2.5 concentrations [11]. From these data we will calculate the mean exposure of each subject to specified air pollution during specific exposure windows.

Other available data on air pollution include ozone in parts per billion, fine particles (number of particles per $cm^3$, average particle diameter in nm, lung-deposited particle surface in $um^2/cm^3$), and temperature and humidity measurements obtained at schools on the day of the study visit using a wireless collection system installed on the roof of the bus in collaboration with the OpenSense team of the ETH (Swiss Federal Institute of Technology in Zurich). The system sent data continuously to a central database at the ETH and sensors were calibrated when the bus passed a reference station.

### Data quality

#### *Parental questionnaires*

The data entry form of the parental questionnaire contained automatic plausibility checks to reduce data entry errors. We did double data entry for a random 10% of the parental questionnaires using the software Epidata. Variables had one to five entry fields. We found a low field error percentage (1%) and thus did not double enter the remaining 90% of questionnaires.

#### *DTG-SBW*

Quality control of DTG-SBW measurements was performed by the field workers on site. A double check of DTG-SBW test results is on-going, to see if the phase III was linear and constituted at least 50% of expired volume [9, 12]. Quality control criteria for DTG-SBW were defined as: 1) no evidence of air leaks as monitored by volume and MM signals, 2) similar flow-volume-loops in pre-test and test breaths, 3) breath volumes of the five tidal pre-test and the test breaths were within 10%, 4) inspiratory peak flow within the by-pass flow.

For signal processing and analyses we used software developed by our group (LungSim, Numerical Modeling, Thalwil, Switzerland) running in (Matlab® R2014a, The Mathworks Inc., Natick, MA, USA) [9]. MM, CO2 and volume signals were aligned in time accounting for different signal rise times [9]. To extract the double-tracer gas signal from MM we subtracted the naturally exhaled CO2 fraction from the MM signal [9]. The corrected MM and CO2 expirograms were plotted against expired volume (Figure S3). The SDTG was computed automatically by linear regression between 65 to 95% of expired volume and under visual control. If required, we manually adjusted volume limits to exclude adjacent tidal phases. Quality criteria were presence of both MM and CO2 phase III over at least 50% of expired volume.

#### *Fractional exhaled nitric oxide (FeNO)*

Required average flow in the plateau was 40-60 ml/s and required duration of exhalation was at least 4 seconds for children younger than 12 years and 6 seconds for children aged 12 years or older. Mean FeNO was calculated using at least two reproducible exhalations with the nitric oxide plateau values within 10% of each other. Field workers checked the quality of the FeNO measurements on site according to ATS/ERS criteria for online FeNO measurement in children [13].

#### *Spirometry flow-volume curves*

In a dedicated workshop, a paediatric pulmonologist and lung function expert (FS) explained the fundamental concepts of paediatric spirometry quality control to all participants. Five pairs of team members assessed and scored by consensus the quality of spirometry curves into “good”, “moderate” or “bad” quality” depending on the presence of hesitation at start of expiration, submaximal effort, forced flow deviations (e.g. cough or glottis closure) within or after the first second, and premature ending of the exhalation within or after the first second. Three paediatric respiratory physicians (FS, CK, JU) assessed all spirometry curves marked as “moderate” or “bad” quality during the first screening and set their own score. If the paediatric respiratory physicians had doubts on the scoring of a spirometry curve, they discussed the curve among them and reached an agreement.

**Table S1:** Characteristics of schools in the canton of Zurich and visited by the *LuftiBus in the school study* (LUIS) at baseline and at one-year follow-up, by degree of urbanisation.

| | **Urbanisation degree** | | | **Total** |
|---|---|---|---|---|
| | **Large urban** | **Small urban** | **Rural** | |
| **Schools in canton of Zurich** | | | | |
| Number of schools: n (%) | 201 (41) | 228 (47) | 61 (12) | 490 |
| Number of classes: n (%) | 2450 (45) | 2602 (48) | 381 (7) | 5433 |
| Number of students: n (%) | 48718 (45) | 51588 (48) | 7333 (7) | 107639 |
| Average number of students per class | 19.9 | 19.8 | 19.2 | 19.8 |
| **Schools visited by LUIS, baseline** | | | | |
| Number of schools: n (%) | 15 (40) | 18 (49) | 4 (11) | 37 |
| Number of classes: n (%) | 149 (39) | 204 (54) | 26 (7) | 379 |
| Number of students: n (%) | 2959 (40) | 3842 (53) | 536 (7) | 7337 |
| Average number of students per class | 19.9 | 18.8 | 20.6 | 19.4 |
| **Schools visited by LUIS, follow-up** | | | | |
| Number of schools: n (%) | 4 (21) | 13 (68) | 2 (11) | 19 |
| Number of classes: n (%) | 45 (23) | 143 (73) | 7 (4) | 195 |
| Number of students: n (%) | 917 (24) | 2679 (72) | 143 (4) | 3739 |
| Average number of students per class | 20.4 | 18.7 | 20.4 | 19.2 |

Calculations are based on aggregated data provided by the Swiss federal statistical office. The number of schools in the canton of Zurich corresponds to the year 2013. Definition of urbanization degree [14]: Large urban area: At least 50% lives in high-density clusters. Small urban area: Less than 50% of the population lives in rural grid cells and less than 50% lives in a high-density cluster. Rural area: More than 50% of the population lives in rural grid cells.

**Table S2:** Number of participants in the LuftiBus in the school (LUIS) study with available information on parental questionnaires (PQ), children's questionnaires (CQ), spirometry (Spiro), and fractional exhaled nitric oxide (FeNO) at baseline and at the one-year follow-up.

| **Available data** | | | | | **Baseline** N=3870 n (%) | **Follow-up** N=655 n (%) |
|---|---|---|---|---|---|---|
| PQ | | | | | 3457 (89) | 629 (96) |
| CQ | | | | | 3546 (92) | 640 (98) |
| Spiro | | | | | 3446 (89) | 614 (94) |
| FeNO | | | | | 3393 (88) | 588 (90) |
| DTG-SBW | | | | | 1795 (46) | 496 (76) |
| PQ | & CQ | | | | 3171 (82) | 614 (94) |
| PQ | & Spiro | | | | 3071 (79) | 589 (90) |
| PQ | & FeNO | | | | 3030 (78) | 565 (86) |
| PQ | & DTG-SBW | | | | 1627 (42) | 472 (72) |
| CQ | & Spiro | | | | 3230 (83) | 610 (93) |
| CQ | & FeNO | | | | 3183 (82) | 584 (89) |
| CQ | & DTG-SBW | | | | 1784 (46) | 492 (75) |
| Spiro | & FeNO | | | | 3165 (82) | 561 (86) |
| Spiro | & DTG-SBW | | | | 1585 (41) | 472 (72) |
| FeNO | & DTG-SBW | | | | 1595 (41) | 451 (69) |
| CQ | & Spiro | & FeNO | | | 2964 (77) | 557 (85) |
| CQ | & Spiro | & DTG-SBW | | | 1579 (41) | 469 (72) |
| CQ | & FeNO | & DTG-SBW | | | 1585 (41) | 448 (68) |
| PQ | & Spiro | & FeNO | | | 2824 (73) | 539 (82) |
| PQ | & Spiro | & DTG-SBW | | | 1429 (37) | 449 (69) |
| PQ | & CQ | & Spiro | | | 2882 (74) | 585 (89) |
| PQ | & CQ | & FeNO | | | 2845 (74) | 561 (86) |
| PQ | & FeNO | & DTG-SBW | | | 1429 (37) | 429 (65) |
| Spiro | & FeNO | & DTG-SBW | | | 1460 (38) | 430 (66) |
| PQ | & CQ | & Spiro | & FeNO | | 2646 (68) | 535 (82) |
| PQ | & CQ | & FeNO | & DTG-SBW | | 1421 (37) | 426 (65) |
| PQ | & Spiro | & FeNO | & DTG-SBW | | 1305 (34) | 409 (62) |
| PQ | & CQ | & FeNO | & DTG-SBW | | 1421 (37) | 426 (65) |
| CQ | & Spiro | & FeNO | & DTG-SBW | | 1454 (38) | 427 (65) |
| PQ | & CQ | & Spiro | & FeNO | & DTG-SBW | 1300 (34) | 406 (62) |

Number of measurements before quality control.

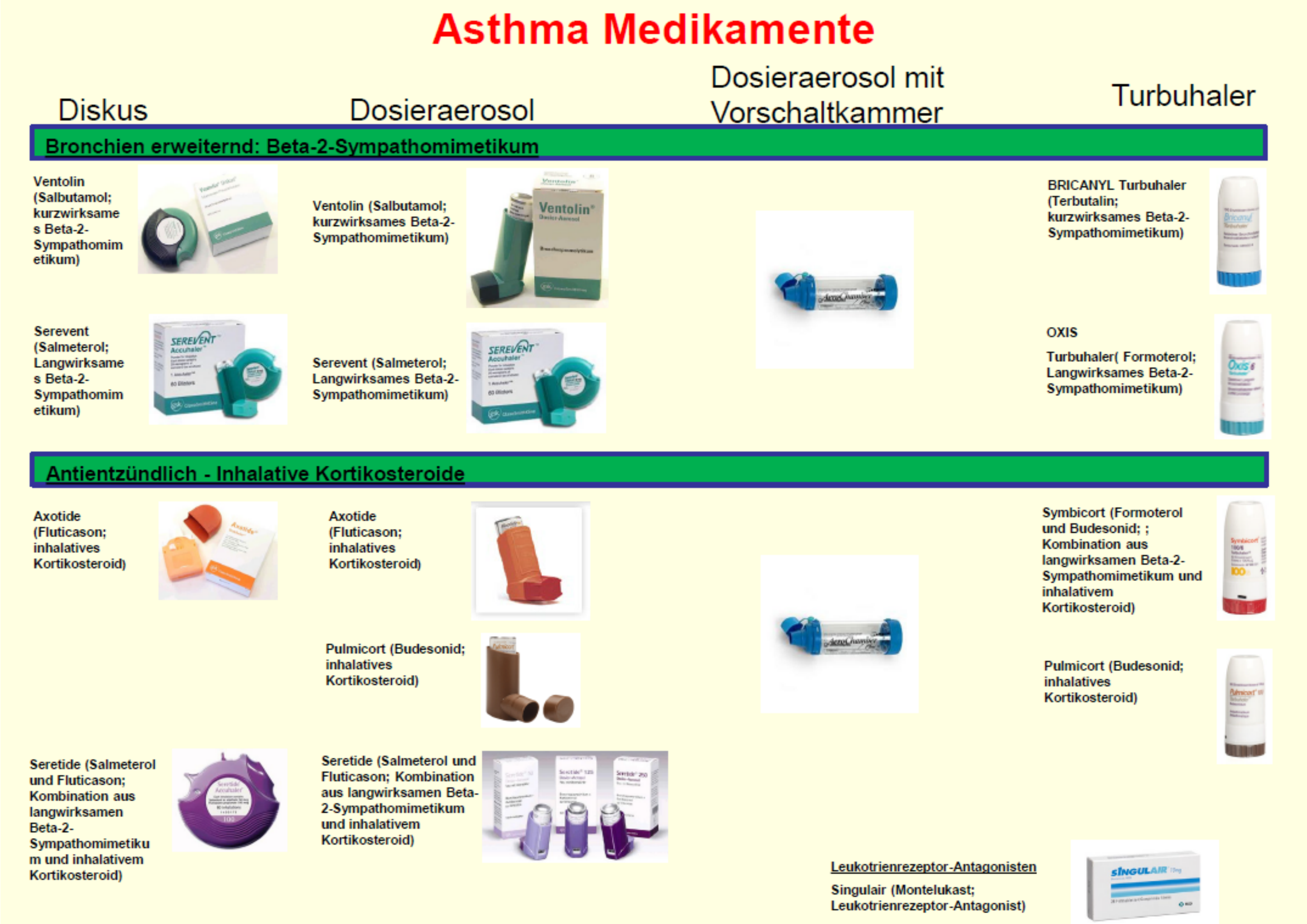


**Figure S1:** Poster of asthma medications used in the *LuftiBus in the school* (LUIS) study to help children answer which medication they had taken the day of the visit (in German).

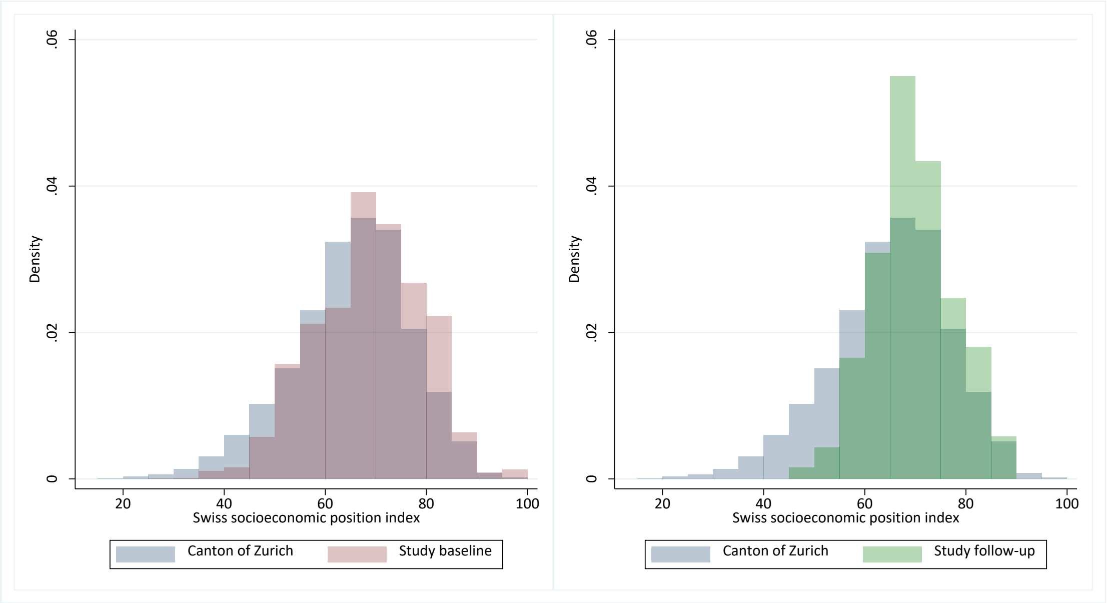


**Figure S2:** Histograms showing the distribution of the Swiss socioeconomic position index for families from the canton of Zurich with at least one child aged 6-17 years living in the household and in the *LuftiBus in the school* (LUIS) study at baseline and one-year follow-up.

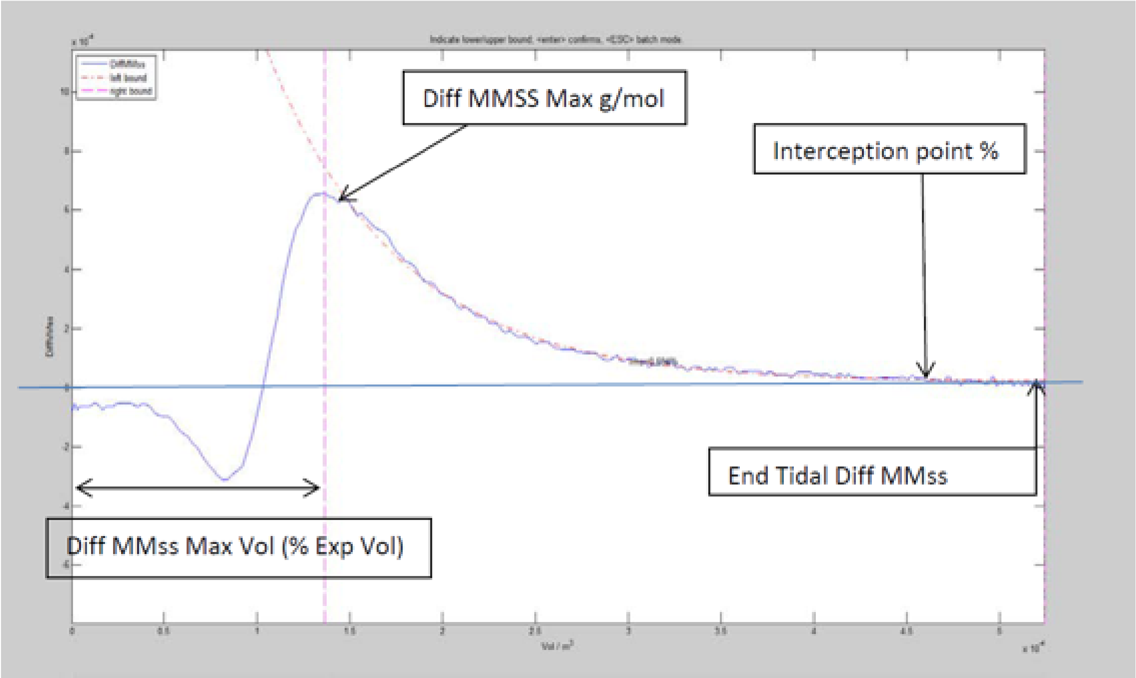


**Figure S3:** Fit lines of double tracer gas single breath washout (DTG-SBW), differences in molar mass (MM) plotted against expired volume.

Abbreviations: MMss = Molar mass signal of the side stream; MMss calc = Calculated molar mass signal of the side stream using measured gas proportion of a normal breath; Diff MMss = MMss – MMss calc; End Tidal Diff MMss: Difference of MMss – MMss calc (in %) at the end of expiration; Interception point (%): % of expired volume where Diff MMss = 0; Diff MMss Max Vol (% Exp Vol): Difference of MMss-MMss calc at the maximum expired volume; Left and right bound= 65-95% of expired volume.
